# Exposotypes in Psychotic Disorders

**DOI:** 10.1101/2025.02.14.25322306

**Authors:** Walid Yassin, Bryan Kromenacker, James B Green, Carol A Tamminga, Elisabetta C. del Re, Pegah Seif, Cuihua Xia, Ney Alliey-Rodriguez, Elliot S Gershon, Brett A Clementz, Godfrey D Pearlson, Sarah S Keedy, Elena I Ivleva, Scott Kristian Hill, Jennifer E McDowell, Matcheri S Keshavan

## Abstract

Psychiatry lags in adopting etiological approaches to diagnosis, prognosis, and outcome prediction compared to the rest of medicine. Etiological factors such as childhood trauma (CHT), substance use (SU), and socioeconomic status (SES) significantly affect psychotic disorder symptoms. This study applied an agnostic clustering approach to identify exposome clusters “Exposotypes (ETs)” and examine their relationship with clinical, cognitive, and functional outcomes. Using data from individuals with psychotic disorders (n=1,350), and controls (n=623), we assessed the relationship between the exposotypes and outcomes. Four exposotypes were identified: ET1 characterized by high CHT and SU; ET2, high CHT; ET3, high SU; ET4, low exposure. Compared to ET4, ET1 demonstrated higher positive and general symptoms, anxiety, depression, impulsivity, and mania; ET2 had higher anxiety, depression, and impulsivity; ET3 had better cognitive and functional outcomes with lower negative symptoms. Intracranial volume was largest in ET3, and smallest in ET2. No group differences in schizophrenia polygenic risk scores were found. The age of onset was 5 years earlier in ET1 than in ET4. These findings provide insight into the complex etiological interplay between trauma, and SU, as well as their unique effects on clinical symptoms, cognition, neurobiology, genetic risk, and functioning.

## Introduction

Psychotic disorders are generally characterized by positive, negative, and other symptoms that can have a significant impact on the well-being of individuals with these conditions. The current diagnostic criteria, primarily based on symptomatology, remain largely subjective and encompass substantial biological and etiological heterogeneity ^1,2^. This heterogeneity has hampered effective treatment for decades. To address such limitations the Bipolar-Schizophrenia Network for Intermediate Phenotypes (B-SNIP) consortium pioneered an alternative approach that considered biological underpinnings of psychosis leading to the creation of the B-SNIP Biotypes (BTs), where BT1 and BT2 were characterized by poor cognition, BT1 by low neural response magnitudes, BT2 by overactive neural responses and poor sensory-motor inhibition, and BT3 was comparable to controls, suggesting paths forward in several domains including diagnosis, prognosis, and treatment approaches ^3–5^.

Etiological approaches in medicine are common and proven to have high value. For example, recognizing the etiological underpinning of coronary artery disease led to the identification of lifestyle risk factors such as diet and exercise in determining prognosis, the formation of the Framingham Risk Score ^6^, and the development of statins ^7^. Despite their value, etiological approaches in psychiatry have been limited. An early example of this approach was conceptualized by Murray et al. who classified schizophrenia into familial and sporadic subtypes ^8^. While genetics plays an important role in psychotic disorders’ etiology, environmental factors such as childhood trauma (CHT), substance use (SU), and socioeconomic status (SES), have become increasingly recognized as important risk factors ^9–11^. These factors can influence the symptom onset and severity, as well as the course of psychosis outcome.

Several previous reports have explored the impact of CHT, SU, and SES as well as their interplay in psychotic disorders ^12–17^. CHT, for example, has been consistently reported to have a negative impact. This includes earlier age of onset ^14,18^, more severe anxiety ^19^, depression ^13,20^, psychotic symptoms ^21,22^, mania ^23^, and impulsivity ^24^. In addition, it is associated with worse cognition ^25^, smaller intracranial (ICV), gray matter (GMV), and hippocampal volume ^14,26,27^, as well as lower overall functioning ^28^. SU has also been studied extensively in psychotic disorders, however, the results are less consistent than those of CHT. Cannabis use (CU), for instance, has been reported to improve anxiety and depression in some studies and worsen them in others ^29–31^. CU has also been associated with worse impulsive behavior, cognition, and psychotic symptoms, though some studies reported reduced symptoms or no change ^30,32–39^. Moreover, reduced or no changes are reported in ICV, GMV, and hippocampal volume ^14,27,40^. Alcohol use (AU) research, on the other hand, demonstrates more consistent findings with reported heightened anxiety, depression, and mania ^16,41–43^. However, there are also reports of larger ICV in AU, with decreased volume in the subcortical regions ^44,45^. In contrast to these inconsistencies, both CU and AU have been regularly linked to an earlier age of psychosis onset ^14,46–49^. SES also impacts psychotic disorder outcomes ^50^. Low SES has been reported to modestly increase the risk of developing psychotic disorders ^50^, anxiety ^51^, depression ^51,52^, and impulsivity ^53–56^. It is also associated with worse cognition ^50,57,58^, and overall functioning ^59^.

CHT, SU, and low SES frequently co-occur and several studies have reported an interaction between these factors. For example, our previous work demonstrated that CHT and CU interact to increase the risk of developing a psychotic condition by lowering the age of onset ^14,60^. Additionally, as outlined in a systematic review, the interaction of CHT with several other substances including AU impacted overall outcomes ^61^. Moreover, low SES can exacerbate the effects of CHT in chronic conditions ^62^ and is linked to both CU and AU ^63,64^. Thus there is a confluence of effects leading to complex interactions between the different environmental factors and their impact on the brain and behavior ^11^.

Given these interactions and others between the myriad environmental factors that can impact an individual throughout their lifespan, understanding which factors share common influences and which operate independently can offer a deeper insight into their overall effects. Classic approaches to risk factor research have mostly focused on delineating the effects of individual factors through direct group comparisons or evaluating the strength of their associations with outcomes ^65,66^. More recent exposome approaches for psychotic disorders have controlled for the covariance among the risk factors themselves. Such an exposeome framework sought to identify the unique weights of the different risk factors that transform their presence or absence into a unidimensional additive composite risk score for an individual ^67,68^. In contrast, an exposotype approach groups similar or co-occurring environmental factors together across multiple exposure dimensions, and can capture more complex interrelations and group dynamics that other methods might overlook. In addition, by evaluating the outcome measures associated with each exposotype group, this complementary framework can uncover unique etiological profiles that may better inform future diagnostic, prognostic, and targeted therapeutic strategies.

This study aimed to identify psychotic disorders exposotypes, as well as characterize them, and their relationship with age of onset, symptoms severity, cognition, ICV, functioning, and genetic risk in psychotic disorders and controls, using a clustering approach. We hypothesized the formation of 4 clusters: CHT, SU, SES, and low-exposure. We also hypothesized that the cluster with CHT would demonstrate worse outcomes across the outcomes evaluated, followed by SU, SES, and low-exposure clusters. We predict that the exposotype characterized as “low-exposure” would have the highest genetic risk. Additionally, regarding the Biotypes, we expect to see BT3 constituting a large portion of the SU ET, while BT1/2 would be distributed in the other Exposotypes (ETs). Lastly, we predicted that there would be larger differences between the exposotype groups in the controls than the psychotic disorders since some of the effects of the exposotypes might overlap with the psychopathology.

## Methods

### Participants

The participants were recruited as part of the B-SNIP2 study, from 5 sites (Athens, GA, Boston, MA, Chicago, IL, Dallas, TX, and Hartford, CT). The sample (n = 1973) comprised 1350 probands and 623 controls. The group with psychotic disorders included those with a DSM-IV diagnosis of schizophrenia (SZ), schizoaffective disorder (SAD), or psychotic bipolar I disorder (BPD) (Table S1). The diagnosis was confirmed using the Structured Clinical Interview for DSM-IV Axis I Disorders (SCID). The patients were clinically stable and in a non-acute symptom state. All diagnostic groups, including controls, were included to ensure an agnostic approach to identifying etiologic clusters. Additionally, this approach helps mitigate potential biases that might arise from focusing solely on participants with psychotic disorders, which could skew cluster formation. All participants provided informed consent. The data is available online at The National Institute of Mental Health Data Archive.

### Assessments

The demographic information of the participants was analyzed for age and gender.

### Socioeconomic status

Socioeconomic status (SES) was evaluated using the Hollingshead scale ^69^. The concept of SES in general is a conglomerate of a wide range of factors including urbanicity, neighborhood characteristics, income, education, etc. Such factors increase the likelihood of multiple poor health outcomes including symptoms experienced in different psychiatric conditions such as psychotic disorders. Here, we use a simplified measure of that in the form of the Hollingshead scale.

### Symptoms and Cognitive Variables

Current symptoms severity in individuals with psychotic disorders were evaluated using the Positive and Negative Syndrome Scale (PANSS), Young Mania Rating Scale (YMRS), Montgomery-Åsberg Depression Rating Scale (MADRS), Clinical Anxiety Scale (CAS), Barratt Impulsiveness Scale (BIS-11), and Birchwood Social Functioning Scale (SFS). Cognitive function was tested using the Brief Assessment of Cognition in Schizophrenia (BACS) battery. For further details, see Tamminga et al. ^3^.

### Childhood trauma

To evaluate history of childhood trauma including emotional abuse, physical abuse, sexual abuse, emotional neglect, and physical neglect, the Childhood Trauma Questionnaire (CTQ) was used ^70^.

### Substance use

Lifetime frequency of cannabis and alcohol use was assessed using the following categories (score 1-6): 1 = none, 2 = 1-4 times, 3 = 5-10 times, 4 = 11-50 times, 5 = 51-100 times, 6 = >100 times. The frequency of cannabis use was assessed before age 15 years, between 15 and 18 years, and after 18 years; and of alcohol use, before 18 and after 18 years.

### Functioning

Functioning was evaluated using the Global Assessment of Functioning (GAF) and the Birchwood Psychosocial Functioning Scale ^71,72^.

### Genetics

Genetic information was extracted from blood DNA processed at the Broad Institute using the Illumina Infinium® Global Screening Array-24 v1.0. (Illumina Inc., San Diego, CA, USA). The array has 688,032 markers mapped to the human genome GRCh37/hg19 reference build. Construction of the trans-ancestry adjusted polygenic risk score (TAPRSs) for schizophrenia included each of the 5 ancestries: European, African, Admixed American, East Asian, and South Asian. More details about the genetic information and the TAPRS can be found elsewhere ^73^.

### Magnetic Resonance Imaging

3 Tesla T1-weights whole brain images were acquired using the Alzheimer’s Disease Neuroimaging Initiative (ADNI) protocol. The images were quality-checked, processed using FreeSurfer v. 7.1, and rigorously evaluated by two independent raters for quality post-processing. The outcomes of interest included total intracranial volume (ICV), total gray matter volume (GMV), and hippocampal volume. The hippocampal volume and GMV were adjusted for ICV.

### Biotypes

Most participants from BSNIP were included in a biotype approach based on electrophysiology and cognition. This work resulted in the determination of 3 Biotypes (BTs) ^4^. BT1 and 2 have poor cognition. BT1 was characterized by low neural response magnitudes, while BT2 by overactive neural responses and poor sensory-motor inhibition. BT3 was comparable to controls^5^.

### Machine learning

#### Preprocessing

Instances with more than 50% of missing data were not included in the analyses. The data were divided into discovery and validation sets with a 50% split and stratified by the study site using the train_test_split functioning in scikit-learn v1.4.2 (https://scikit-learn.org/). The site stratification allowed each sample to have representation from various geographic areas. The input variables for the machine learning included CHT using the CTQ, substance use, and socioeconomic status (participant and parent) (See *Methods* section for further details about the variables).

All participants, including individuals with psychotic disorders and controls, were included. The discovery data was imputed using KNNImputer, part of Scikit Learn v1.4.2 (https://scikit-learn.org/), the imputation of the discovery data was then applied to validation data (<2% in both datasets) using the imputer fit/transform functions. Site correction was performed using ComBat Family of Harmonization Methods (https://github.com/andy1764/ComBatFamily) in RStudio v4.3.2. This was also applied to the validation set, an option available with this new version of Combat. After this step, the data were standardized using scikit learn’s preprocessing function “StandardScalar”, and was applied to the validation data using the fit-transform function. Principal component analysis (PCA), part of the decomposition package in scikit learn, was used as the last step before clustering. This step was also used on the discovery data and applied to the validation data.

#### Clustering

Clustering was conducted using clValid v0.7 in R ^76^. We utilized k means, AGglomerative NESting (Agnes), DIvisive ANAlysis Clustering (DIANA), and Partition Around Medoids (PAM), iterating over their available parameters. We chose the best-performing algorithm and number of clusters based on the Silhouette score. Cluster membership was extracted from that algorithm, and used for further analysis. This hierarchical clustering approach has been used in several studies in CHR before ^77–79^.

#### Cluster Validation

The discovery clusters were validated in two ways ^80^. The first approach was *Methods-based* validation. Here, the principal components from the validation dataset were evaluated using the same algorithm, parameters, and number of clusters that were optimal for the discovery dataset. The results of these runs were then compared using their Silhouette score, where a similar score indicates good validation ^80^. In the second approach, the *Results-based* validation, the centroids of the discovery dataset were obtained and the instances in the validation dataset were mapped on them where the proximity of instances in the validation dataset to the centroids determines their cluster membership ^80^. A random forest classifier was used from sklearn using the cluster membership as a label and the instances’ PCs as input variables from the discovery data as the training set. This trained algorithm was then used to classify the validation data. Accuracy was used to report the results. The precision, recall, f1-score, and confusion matrix were also provided.

#### Statistical analyses

Demographic characteristics of the clusters were delineated using chi-square and ANOVA where appropriate. Initial comparison of clusters was done using the Kruskal-Wallis test and post-hoc pairwise using Dunn’s test given the non-normality of the data. The outcome comparisons were conducted for both the clinical and control samples using ANOVA and t-tests. All outcome comparison p-values were adjusted using the False Discovery Rate (FDR) method to *q* values. The percentages were calculated within each group for all categorical variables. Critical values of the adjusted residuals from the chi-square analyses were used to identify significant differences between cluster and other grouping variables. Z scores were calculated by subtracting the mean from a raw score and dividing it by the standard deviation.

## Results

### Machine learning

#### Clustering

Using the discovery data, the clustering analysis demonstrated that DIANA was the optimal algorithm resulting in 4 clusters with good separability indicated by the silhouette score of 0.66; Cluster 1 or exposotype 1 (ET1) characterized by high CHT, SU, and moderate SES, exposotype 2 (ET2) by high trauma and moderate SES, exposotype 3 (ET3) by substance use and moderate SES, and exposotype 4 (ET4) by low overall exposure, i.e., low CHT, low SU, and moderate SES, cluster in one group. The ET4 group was designated as our “exposome control” (Figure 1a-c, Table 1). The detailed clustering results can be found in Table 1.

**Figure 1:**
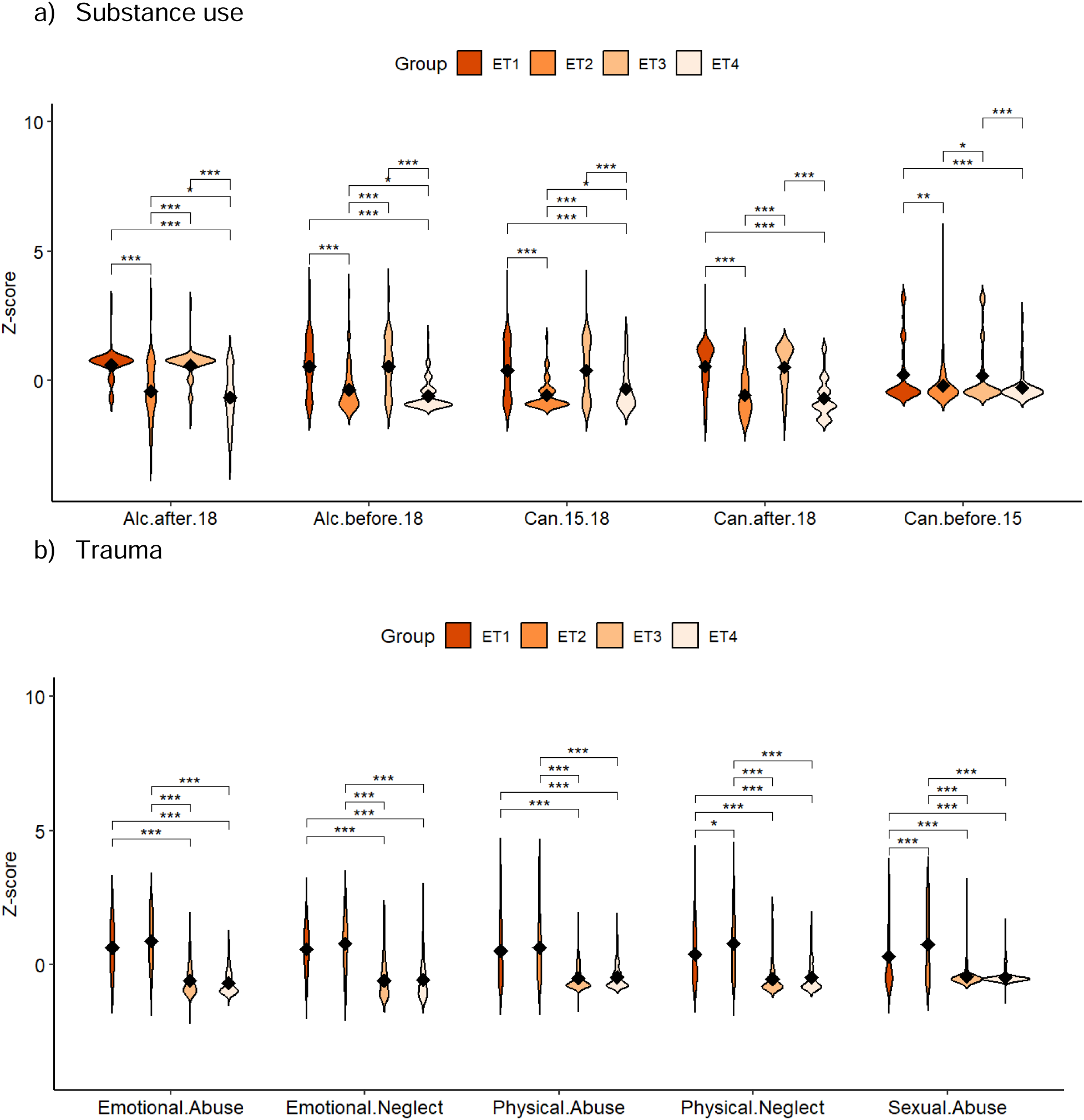
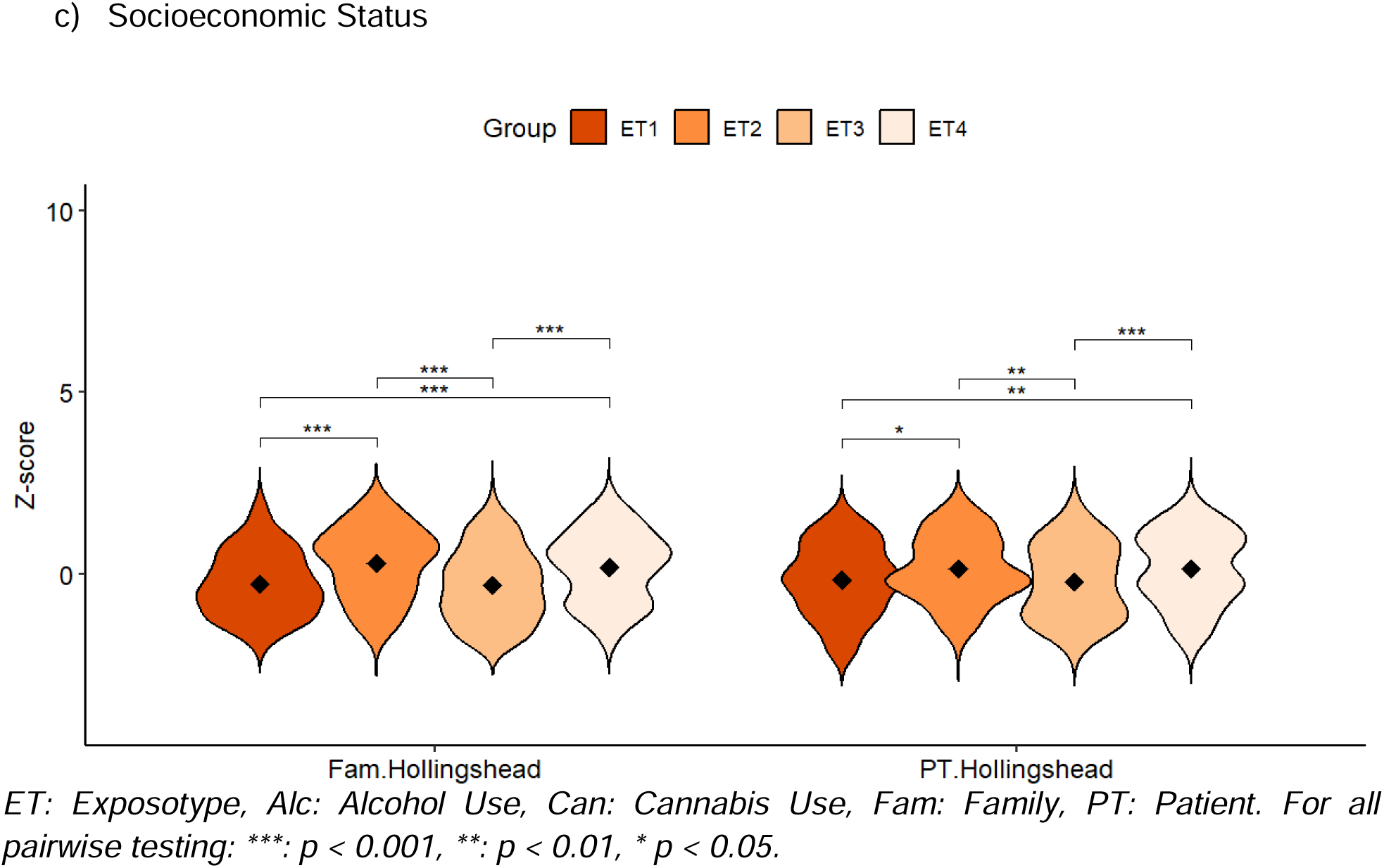

**Table 1:** Exposotype Identification.

|  | Outcome | ET1<br>(□) | ET2<br>(□) | ET3<br>(⊗) | ET4<br>(Δ) |  |  |  |  |
| --- | --- | --- | --- | --- | --- | --- | --- | --- | --- |
| | | M (SD) | | | | H | DF | $\eta^2G$ | $p$ |
| Substance | Alcohol after 18 | 5.77<br>(0.71) | 4.39<br>(1.43) | 5.75<br>(0.67) | 4.04<br>(1.4) | 256.68 | 3 | 0.37 | □□⊗Δ = ***<br>□□ = ***<br>□Δ = ***<br>□⊗ = ***<br>□Δ = *<br>⊗Δ = *** |
|  | Alcohol before 18 | 3.75<br>(1.91) | 2.06<br>(1.47) | 3.73<br>(1.88) | 1.61<br>(0.99) | 188.81 | 3 | 0.258 | □□⊗Δ = ***<br>□□ = ***<br>□Δ = ***<br>□⊗ = ***<br>□Δ = *<br>⊗Δ = *** |
|  | Cannabis between 15 - 18 | 3.51<br>(2) | 1.68<br>(1.1) | 3.55<br>(2.1) | 2.16<br>(1.51) | 86.36 | 3 | 0.115 | □□⊗Δ = ***<br>□□ = ***<br>□Δ = ***<br>□⊗ = ***<br>□Δ = *<br>⊗Δ = *** |
| | Cannabis after 18 | 4.78<br>(1.55) | 2.76<br>(1.49) | 4.73<br>(1.5) | 2.52<br>(1.35) | 175.08 | 3 | 0.23 | $\square\square\otimes\Delta =$<br>***<br>$\square\square =$ ***<br>$\square\Delta =$ ***<br>$\square\otimes =$ ***<br>$\otimes\Delta =$ *** |
| | Cannabis before 15 | 1.94<br>(1.6) | 1.36<br>(1.01) | 1.89<br>(1.64) | 1.24<br>(0.77) | 21.13 | 3 | 0.031 | $\square\square\otimes\Delta =$<br>***<br>$\square\square =$ **<br>$\square\Delta =$ ***<br>$\square\otimes =$ *<br>$\otimes\Delta =$ *** |
| Trauma | Emotional abuse | 14.38<br>(4.73) | 15.65<br>(5.4) | 7.38<br>(2.98) | 6.89<br>(2.43) | 378.82 | 3 | 0.497 | $\square\square\otimes\Delta =$<br>***<br>$\square\otimes =$ ***<br>$\square\Delta =$ ***<br>$\square\otimes =$ ***<br>$\square\Delta =$ *** |
| | Emotional neglect | 14.29<br>(4.39) | 15.45<br>(4.99) | 8.21<br>(3.1) | 8.39<br>(3.58) | 293.5 | 3 | 0.382 | $\square\square\otimes\Delta =$<br>***<br>$\square\otimes =$ ***<br>$\square\Delta =$ ***<br>$\square\otimes =$ ***<br>$\square\Delta =$ *** |
| | Physical abuse | 10.94<br>(5.1) | 11.48<br>(5.29) | 6.24<br>(2) | 6.47<br>(2.01) | 232.51 | 3 | 0.312 | $\square\square\otimes\Delta =$<br>***<br>$\square\otimes =$ ***<br>$\square\Delta =$ ***<br>$\square\otimes =$ ***<br>$\square\Delta =$ *** |
| | Physical neglect | 9.5<br>(3.53) | 11<br>(4.21) | 6.14<br>(1.95) | 6.33<br>(1.97) | 260.4 | 3 | 0.351 | $\square\square\otimes\Delta =$<br>***<br>$\square\square = *$<br>$\square\otimes = ***$<br>$\square\Delta = ***$<br>$\square\otimes = ***$<br>$\square\Delta = ***$ |
| | Sexual abuse | 9.97<br>(6.69) | 12.67<br>(7.43) | 5.61<br>(2.33) | 5.45<br>(1.52) | 211.48 | 3 | 0.359 | $\square\square\otimes\Delta =$<br>***<br>$\square\square = ***$<br>$\square\otimes = ***$<br>$\square\Delta = ***$<br>$\square\otimes = ***$<br>$\square\Delta = ***$ |
| Socioeconomic status | Family Hollingshead | 36.24<br>(14.21) | 45.14<br>(15.57) | 35.7<br>(15.95) | 43.46<br>(15.47) | 47.71 | 3 | 0.062 | $\square\square\otimes\Delta =$<br>***<br>$\square\square = ***$<br>$\square\Delta = ***$<br>$\square\otimes = ***$<br>$\otimes\Delta = ***$ |
| | Patient Hollingshead | 40.35<br>(14.86) | 44.92<br>(13.9) | 39.7<br>(15.46) | 45.04<br>(15.29) | 18.48 | 3 | 0.024 | $\square\square\otimes\Delta =$<br>***<br>$\square\square = *$<br>$\square\Delta = **$<br>$\square\otimes = **$<br>$\otimes\Delta = ***$ |

ET: Exposotype, □ : ET1, □ : ET2, $\otimes$ : ET3, $\Delta$ :ET4, M: Mean, SD: Standard Deviation, H: Kruskal Wallis test statistic, DF: Degrees of Freedom, $\eta^2$ G: Generalized Eta Squared

#### Validation

The discovery dataset demonstrated a similar silhouette score of 0.66 to the validation of 0.67. The validation method involving the supervised classification demonstrated an accuracy of 92% (Table S2).

#### Demographic data

The exposotypes differed in age; ET1 and ET2 tended to be 2-3 years older on average than ET3 and ET4. ET2 had noticeably more females while ET3 had more males (Table 2).

**Table 2:** Demographics of discovery sample (n = 772)

| | ET1<br>(N = 152) | ET2<br>(N = 186) | ET3<br>(N = 194) | ET4<br>(N = 240) | Total<br>(N = 772) | DF | $\chi^2$/F | *p* |
| --- | --- | --- | --- | --- | --- | --- | --- | --- |
| Age<br>(S.D.) | 38.63<br>(11.53) | 39.21<br>(12.22) | 36.65<br>(11.98) | 36.01<br>(12.83) | 37.46<br>(12.27) | 3 | 3.13 | 0.025 |
| Sex |  |  |  |  |  | 3 | 46.03 | < 0.001 |
| Male | 82<br>(53.9%) | 55<br>(29.6%) | 122<br>(62.9%) | 105<br>(44.1%) | 364<br>(47.3%) |  |  |  |
| Female | 70<br>(46.1%) | 131<br>(70.4%) | 72<br>(37.1%) | 133<br>(55.9%) | 406<br>(52.7%) |  |  |  |

*ET: Exposotype, S.D.: Standard Deviation, DF: Degrees of Freedom, $\chi^2$ : Chi-square test statistic, F: Anova Test statistic.*

#### Clinical

In the psychotic disorders, those in ET1 showed the highest scores on the PANSS-Positive and PASS-General Psychopathology subscales compared to the other groups, with the severity of positive (PS) and general psychopathology symptoms (GS) significantly differing from ET3 (PS: *q* < 0.001; GS: *q* < 0.01) and ET4 (PS: *q* < 0.01; GS: *q* < 0.05). ET2 also demonstrated significantly higher PS and GS than ET3 (ps: *q* < 0.05; GS: *q* < 0.05). ET4 did not differ from ET2 and ET3. Regarding the PANSS-Negative subscale score, ET1 (*q* < 0.05), ET2 (*q* < 0.05), and ET4 (*q* < 0.01) were significantly higher than ET3. ET1 and ET2 both demonstrated significantly higher levels of anxiety and impulsivity than ET3 (*q* < 0.05) and ET4 (*q* < 0.05), while ET1 and ET2 had significantly higher depression than ET4 (*q* <0.05). ET1 had significantly higher mania scores than ET2 (*q* < 0.05), ET3 (*q* < 0.001), and ET4 (*q* < 0.01) (Table 3, Figure 2a-c). In the control sample, impulsivity, the only measure available for them, was higher in ET1 and ET2 compared to ET3 and ET4 (*q* <0.05) (Table S3).

**Figure 2a-c:**
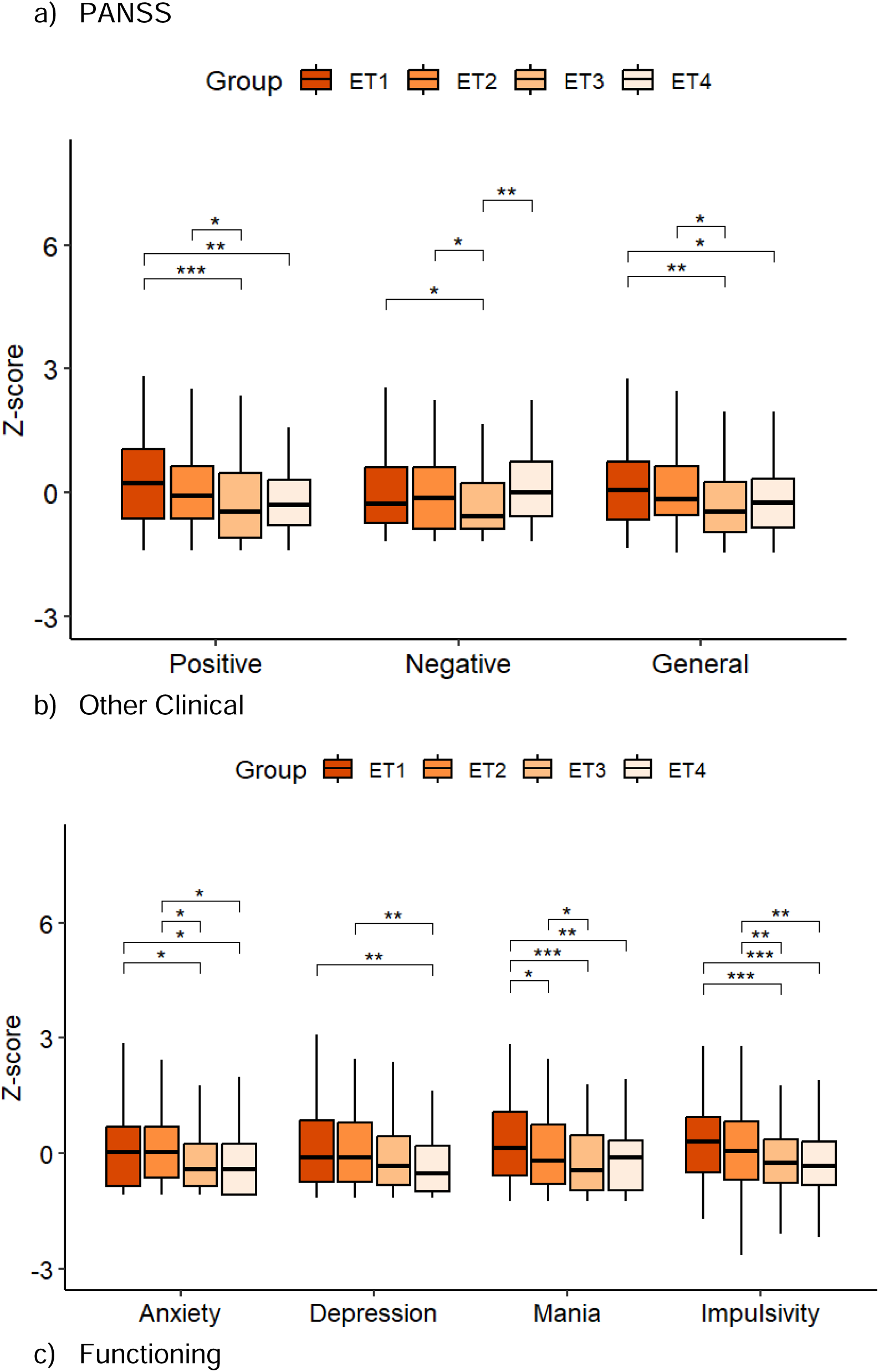
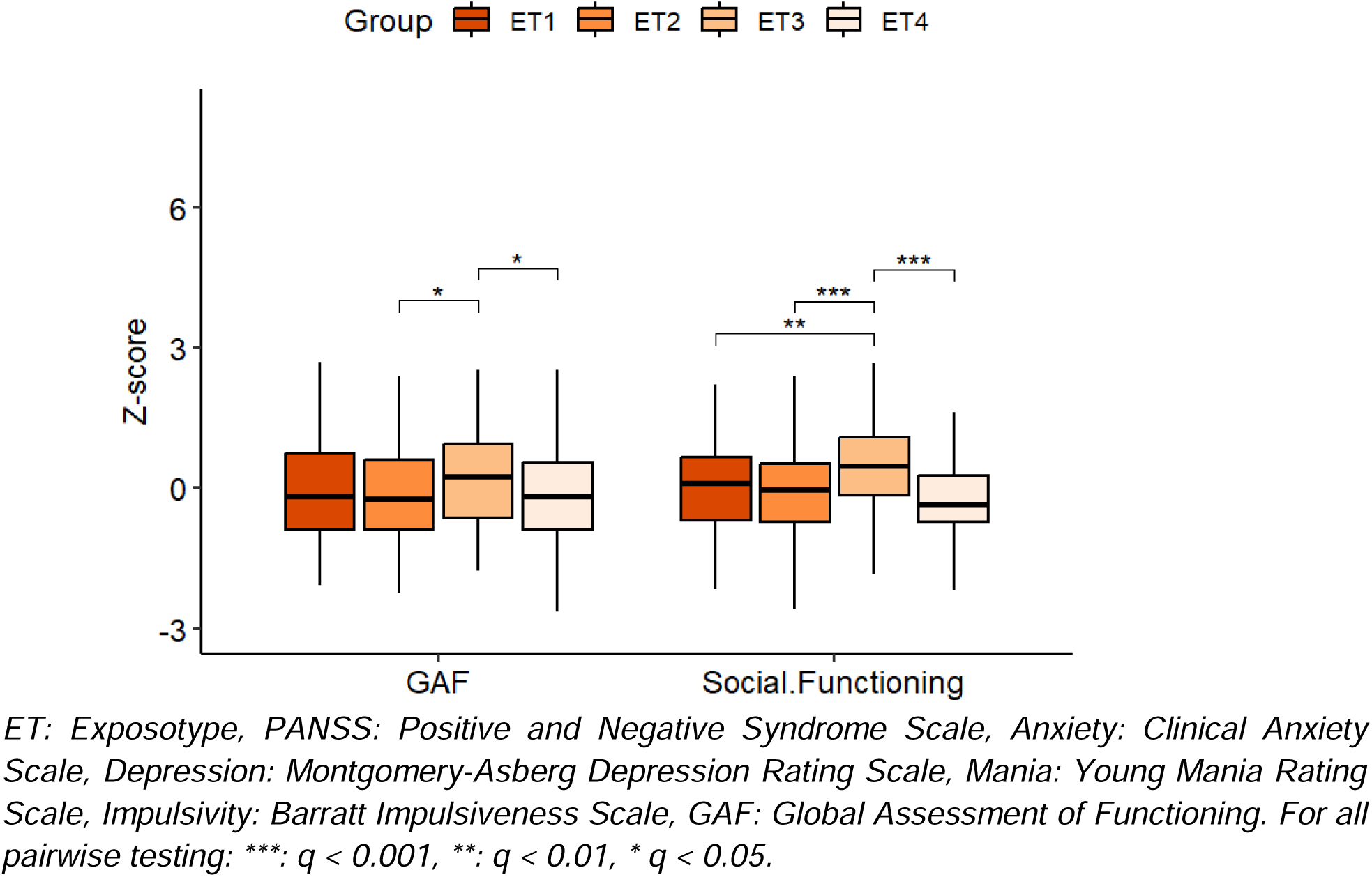
Clinical outcomes and pairwise comparisons for psychotic disorders sample.

**Table 3:** Clinical outcomes and exposotypes for psychotic disorders sample.

|  | Outcome | ET1<br>(□) | ET2<br>(□) | ET3<br>(⊗) | ET4<br>(Δ) |  |  |  |  |
| --- | --- | --- | --- | --- | --- | --- | --- | --- | --- |
| | | M (SD) | | | | DF | F | $\eta^2 G$ | $q$ |
| PANSS | Positive | 17.89<br>(7.23) | 16.42<br>(6.12) | 14.33<br>(6) | 14.9<br>(5.46) | 3 | 7.06 | 0.045 | □□⊗Δ<br>= ***<br>□⊗ = ***<br>□Δ = **<br>□⊗ = * |
|  | Negative | 15.62<br>(7.55) | 14.98<br>(7.26) | 12.98<br>(5.4) | 15.88<br>(5.8) | 3 | 4.03 | 0.026 | □□⊗Δ<br>= **<br>□⊗ = *<br>□⊗ = *<br>⊗Δ = ** |
|  | General | 33<br>(12.1) | 31.29<br>(9.7) | 27.89<br>(8.4) | 29.35<br>(8.64) | 3 | 5.5 | 0.036 | □□⊗Δ<br>= **<br>□⊗ = **<br>□Δ = *<br>□⊗ = * |
| Clinical | CAS | 5.64<br>(4.98) | 5.54<br>(4.49) | 4.08<br>(4.3) | 4<br>(4.16) | 3 | 4.42 | 0.029 | □□⊗Δ<br>= **<br>□⊗ = *<br>□Δ = *<br>□⊗ = *<br>□Δ = * |
| | MADRS | 12.49<br>(10.19<br>) | 12.4<br>(10.17<br>) | 10.59<br>(9.52) | 8.19<br>(7.95) | 3 | 4.89 | 0.031 | $\square\square\otimes\Delta$<br>$= **$<br>$\square\Delta = **$<br>$\square\Delta = **$ |
| | YMRS | 11.84<br>(8.79) | 9.43<br>(7.4) | 7.48<br>(6.22) | 8.39<br>(7.07) | 3 | 6.7 | 0.043 | $\square\square\otimes\Delta$<br>$= ***$<br>$\square\square = *$<br>$\square\otimes =$<br>$***$<br>$\square\Delta = **$<br>$\square\otimes = *$ |
| | BIS-11 | 72.45<br>(12.47<br>) | 70.5<br>(13.63<br>) | 66.34<br>(10.69<br>) | 65.87<br>(11.12<br>) | 3 | 7.98 | 0.048 | $\square\square\otimes\Delta$<br>$= ***$<br>$\square\otimes =$<br>$***$<br>$\square\Delta = ***$<br>$\square\otimes = **$<br>$\square\Delta = **$ |
| Functioning | GAF | 53.14<br>(13.47<br>) | 51.96<br>(11.43<br>) | 56.24<br>(12.77<br>) | 51.62<br>(12.35<br>) | 3 | 3.22 | 0.020 | $\square\square\otimes\Delta$<br>$= *$<br>$\square\otimes = *$<br>$\otimes\Delta = *$ |
| | Birchwood: Social Functioning | 121.8<br>(25.94<br>) | 118.6<br>4<br>(26.19<br>) | 132.4<br>3<br>(24.02<br>) | 115.1<br>8<br>(23.64<br>) | 3 | 9.6 | 0.058 | $\square\square\otimes\Delta$<br>$= ***$<br>$\square\otimes = **$<br>$\square\otimes =$<br>$***$<br>$\otimes\Delta =$ |
|  |  |  |  |  |  |  |  |  | *** |

*ET: Exposotype, □: ET1, □: ET2, ⊗: ET3, Δ:ET4, M: Mean, SD: Standard Deviation, F: ANOVA test statistic, DF: Degrees of Freedom, $r^2$ G: Generalized Eta Squared, q: False-discovery rate converted p-value, PANSS: Positive and Negative Syndrome Scale, CAS: Clinical Anxiety Scale, MADRS: Montgomery-Asberg Depression Rating Scale, YMRS: Young Mania Rating Scale, BIS-11, Barratt Impulsiveness Scale, GAF: Global Assessment of Functioning.*
*For all pairwise testing: \*\*\*: $q < 0.001$ , \*\*: $q < 0.01$ , \* $q < 0.05$ .*

#### Functioning

In the psychotic disorders group, ET3 had significantly higher GAF scores compared to ET2 (*q* < 0.05) and ET4 (*q* < 0.05), as well as significantly higher SFS-based social functioning than ET1 (*q* < 0.01), ET2 (*q* < 0.001), and ET4 (*q* < 0.001). There were no significant differences between the exposotypes for the control group (Table 3).

#### Cognition

In the psychotic disorders group, the BACS score was significantly higher in ET3 compared to ET2 (*q* < 0.01) and ET4 (*q* < 0.001). ET1 and ET2 had BACS composite scores intermediate between ET3 and ET4, whereas ET1 had higher scores than ET2 (Table 4), and ET4 had the lowest scores, significantly lower than ET1 (*q* < 0.001). Additionally, cognitive performance was significantly higher in ET3 than in ET4 in controls (Table S3).

**Table 4:** Exposotypes and differences in Diagnosis, Biotype, Age of onset, Cognition, and PRS for psychotic disorders sample.

| Outcome | ET1 (□) | ET2 (□) | ET3 (⊗) | ET4 (Δ) | DF | F/X<sup>2</sup> | η<sup>2</sup>G/V | q |
| --- | --- | --- | --- | --- | --- | --- | --- | --- |
| Diagnosis |  |  |  |  | 6 | 38.29 | 0.192 | < 0.001*** |
| BPD | 30<br>(30.93%) | 21<br>(21.65%) | 32<br>(32.99%) | 14<br>(14.43%) |  |  |  |  |
| SAD | 51<br>(24.64%) | 79<br>(38.16%) | 38<br>(18.36%) | 39<br>(18.84%) |  |  |  |  |
| SZ | 40<br>(18.6%) | 51<br>(23.72%) | 47<br>(21.86%) | 77<br>(35.81%) |  |  |  |  |
| Biotype |  |  |  |  | 6 | 19.33 | 0.136 | < 0.01** |
| BT1 | 25<br>(16.34%) | 42<br>(27.45%) | 43<br>(28.1%) | 43<br>(28.1%) |  |  |  |  |
| BT2 | 50<br>(28.24%) | 55<br>(31.07%) | 23<br>(12.99%) | 49<br>(27.68%) |  |  |  |  |
| BT3 | 46<br>(24.34%) | 54<br>(28.57%) | 51<br>(26.98%) | 38<br>(20.11%) |  |  |  |  |
| Age of onset | 16.88<br>(7.15) | 17.82<br>(9.29) | 18.4<br>(6.92) | 21.08<br>(8.8) | 3 | 5.639 | 0.034 | □□⊗Δ<br>= **<br>□Δ = ***<br>□Δ = *<br>⊗Δ = * |
| BACS | -1.39<br>(1.27) | -1.66<br>(1.37) | -1.08<br>(1.32) | -2.05<br>(1.34) | 3 | 11.3 | 0.065 | □□⊗Δ<br>= ***<br>□Δ = *** |
| | | | | | | | | $\square \otimes = **$<br>$\otimes \Delta =$<br>*** |
| TAPRS | 0.23<br>(0.46) | 0.22<br>(0.45) | 0.2 (0.5) | 0.12<br>(0.44) | 3 | 1.011 | 0.008 | 0.388 |

ET: Exposotype, □ : ET1, □ : ET2, $\otimes$ : ET3, $\Delta$ : ET4, DF: Degrees of Freedom, F: Anova Test Statistic, $\chi^2$ : Chi-squared test statistic, $\eta^2$ G, Generalized Eta Squared Effect Size, V: Cramer's-V Effect Size, q: False Discovery Rate adjusted p-value, BPD: Bipolar Disorder with Psychotic Features, SAD: Schizoaffective Disorder, SZ: Schizophrenia, BT: Biotype, BACS: Brief Assessment of Cognition in Schizophrenia, Trans-Ancestry Adjusted Polygenic Risk Score for Schizophrenia, For all pairwise testing: \*\*\*: $q < 0.001$ , \*\*: $q < 0.01$ , \* $q < 0.05$ .

#### Age of Onset

Within the psychotic disorders, ET1 (Mean (M): 16.8 years) showed the earliest age of onset followed by ET2 (m = 17.8 years) and ET3 (m = 18.4 years); the latest onset was in ET4 (21.1 years), and ET4 was significantly higher than ET1 (*q* < 0.001), ET2 (p < 0.01), and ET3 (*q* < 0.001). However, ET1, ET2, and ET3 were not significantly different from each other (Table 4).

#### Trans-Ancestry Polygenic Risk Scores

No significant differences were observed between exposotype groups in the ancestry-adjusted PRS (Table 4).

#### MRI-Based Volumetric Outcomes

In the psychotic disorders sample, ET2 had significantly lower ICV than ET3 (*q* < 0.001), and ET1 (*q* < 0.001). ET4 also had significantly lower ICV than ET3 (*q* < 0.05). For the control sample, ET3 was significantly higher in ICV than ET2 (*q* < 0.05) and ET4 (*q* < 0.01), and the right hippocampal volume in ET3 was lower than in ET4 (*q* < 0.01). No other differences were observed (Figure S1-S2, Table S4-S5).

#### Diagnosis, Biotypes, and exposotypes

ET membership was significantly different across DSM-IV diagnostic (□^2^=38.29, *q* < 0.001) and Biotype (□^2^=19.33, *q* < 0.01) groups. In the psychotic disorder group, BPD cases were more prevalent in ET3 (32.99%) and least prevalent in ET4 (14.43%), SAD most prevalent in ET2 (38.16%), and SZ most prevalent in ET4 (35.81%) and, least, in ET1 (18.6%). Among the Biotypes, BT1 was least prevalent in ET1 (16.34%), BT2 was least likely found in ET3 (12.99%), and BT3 was least in ET4 (20.11%) (Figure S3, Table 4). The controls were highly represented in ET3 (30.43%) and ET4 (43.48%) (Table S6).

## Discussion

This is the first study to examine associations between the environmental factors, i.e., CHT, SU (alcohol and cannabis use), and SES, and clinical, cognitive, brain structure, and genetic characteristics in psychotic disorders using a clustering approach. Even though there were four clusters produced, as hypothesized, the nature of the clusters differed from what we initially expected. Despite the significant differences we observed in SES between the groups, the means of these groups remain in the same categorical classification defined by Hollingshead as “class 3” having a score between 32 and 47. Thus we did not stress their contribution when comparing the resulting clusters. Our findings demonstrate higher PANSS positive and general psychopathology symptoms and mania in ET1 compared to those in ET4. We also reported higher anxiety, depression, and impulsivity in ET1, but these were also higher in ET2 than in ET4. Additionally, cognition was higher in ET1, and ET3, than in ET4. ET3 also showed better functioning and lower negative symptoms than ET4. The data available for the control group broadly mirrored those of the probands, with higher cognition in ET3 and worse clinical symptoms in ET2. However, there were no differences in general and social functioning between the groups.

The results of ET1 are consistent with previous literature including our work demonstrating an interaction between these exposotypes and their impact on psychotic symptoms ^14,61^. One postulate could be the relevance to dopamine. Research on trauma demonstrated its ability to increase dopamine dysregulation, predisposing individuals to psychotic symptoms ^81^. This dysregulation can be worsened by substance use which can also affect dopamine. Cannabis and alcohol use can also impact dopamine release leading to worsened psychotic symptoms ^82–84^. The *final common pathway* dopamine hypothesis of schizophrenia links etiology, including CHT, SU, and genetic risk with an increase in presynaptic striatal dopaminergic function ^85^, which in turn could lead to aberrant salience to neutral stimuli that can lead to positive symptoms ^82^. The collective impact of the different exposotypes, such as in ET1, can potentially lead to an exaggerated hyperdopaminergia that might manifest as more severe positive symptoms than the other exposotype groups (ET2/3/4) ^85^. Even though such a relationship cannot be ascertained from the current data, it could be one potential explanation for the lack of differences in positive symptoms between ET2/3 and ET4, but observing a significantly higher symptom severity in ET1 compared to ET4. Indeed, several other neurotransmitters, such as glutamate, also have a role ^86^. Another postulate could be a mechanism taking place in the hippocampus. Evidence from del Re et al. and others demonstrated that CHT and cannabis use interact to lower the AOO of psychosis and that this decrease was associated with higher positive symptoms ^14,87^. This interaction, especially between CHT and SU, is not limited to positive symptoms, but other psychotic symptoms as well including general symptoms ^88^, which could also explain the higher general symptom scores seen in ET1. Negative symptoms did not seem to be different from ET4 in this cohort but were less pronounced in those using substances. This is in alignment with a study by Sabe et al demonstrating that cannabis-using individuals with schizophrenia might be less susceptible to the development of negative symptoms ^38^. Additionally, individuals in the prodromal phase who use substances compared to those who don’t, report less severe negative symptoms ^89,90^ which could potentially explain the lower scores seen in ET3. Moreover, It is also possible that individuals with less negative symptoms are more likely to seek and have more access to substances.

Intriguingly, ET3 did not demonstrate significant impairment overall compared to ET4. On the contrary, it showed a slightly better outcome. The self-medication hypothesis could be one possible explanation. According to that hypothesis, people with schizophrenia might seek to use substances to alleviate their symptoms and medication side effects ^91^. However, as previously mentioned, it is also possible that those with better cognition and functioning are more successful at having access to substances.

Despite their pronounced symptom burden, individuals in ET2 did not exhibit significant cognitive or functional impairments compared to the other groups. This pattern could suggest a potential dissociation between symptom severity and cognitive/functional outcomes, indicating that the detrimental effects of trauma may manifest primarily at the clinical level. These results could potentially mean that other mediating or moderating factors, mechanisms, or compensatory cognitive strategies are at play. Alternatively, it is plausible that this group had access to protective factors, such as higher baseline cognitive reserve which is associated with better functioning ^79^.

Similar to prior literature, those in ET2 showed lower ICV ^26,92,93^, while those in ET3 showed higher ICV which could potentially be driven by the alcohol group ^44,45^. The lack of differences in the hippocampus could be attributed to the impact of psychotic disorders ^94^, where it does not change further with additional environmental insults. We do however see a decrease in the ET3 controls ^95,96^ suggesting a negative impact of SU on the hippocampus in healthy individuals.

Our observation of lower AOO in ET1 and 2 aligns with our previous work, demonstrating that CHT and SU interact to decrease the AOO ^14^. However, evidence from CHT and SU, independently, has shown an effect leading to younger AOO likely due to their impact on the neurodevelopmental trajectories. For example, children experiencing physical trauma are likely to report psychotic symptoms at around age 12 independent of the genetic liability for psychosis^13^.

The findings seen in the clinical and cognitive data of the control group also follow our secondary hypothesis. However, the lack of differences in functioning we observed in psychotic disorders could indicate that controls could have higher functioning independent of SU. Nonetheless, the findings are inconclusive.

The participants’ distribution in the ETs is quite interesting. Affective psychotic disorders, such as SAD and BP were least prevalent in ET4 while non-affective psychotic disorders, SZ here, were most prevalent in that ET group. However, the TAPRS did not differ between the groups; thus, we could not conclude that genetic risk differs between subgroups defined by environmental exposure from the current findings. Nonetheless, the lack of significant genetic risk differences between the groups provides less genetic bias when comparing the exposotypes. On the other hand, BT2, characterized by an over-reactive neural response, was the least prevalent in ET3. It is possible that those with heightened neural reactivity are more sensitive to substance use and that would deter individuals in BT2 from using substances. It could also be the case that substances exacerbate the difficulties in sensory motor inhibition which leads individuals to stay away from using substances. BT3 is mostly represented in the ET2/3 which speaks to its previously hypothesized etiological nature ^3,4^.

### Conclusions

The exposotype approach to disorder classification used in this study is a unique departure from both phenotypic and Biotype approaches, yet highly complementary, in psychotic disorders but not foreign to the rest of medicine. The approach highlights subgroups with specific exposotype signatures that could benefit from tailored intervention. A biotype-informed therapeutic approach supplemented by an exposotype one could potentially have high value as treatment would go beyond the focus on current impairment and would reduce the risk that continues to contribute to the symptom progression and severity. These findings provide insight into the complex etiological interplay between trauma, substance use, and their unique effects on clinical symptoms, cognition, neurobiology, and functioning. Future research exploring the causal association between the exposotypes and outcome measures such as the ones explored in this study as well as the relationship with the diagnostic and biotype group would be highly valuable to pursue.

### Limitations

The non-psychotic control group did not have all the outcome measures available for the proband in the BSNIP dataset, thus no comparisons were performed on those measures. ET4 is considered here to have low etiological impact only related to SU, CHT, and SES, other environmental factors that have not been assessed in this study can still have an effect. It is also important to acknowledge that the current symptom measures are limited in that these are treated patients, i.e., most are on medications. Additionally, the SU measures used in this study are limited in how well they capture use. No causal relationship can be ascertained in the findings of this study.

## Acknowledgments

We acknowledge the contribution of our colleague Dr. John A. Sweeney to the BSNIP data collection. This work was supported by the National Institute of Mental Health (R21MH133001 to Dr. Yassin). Additional support was provided in part by NIMH grants MH-077851 (to CAT), MH-077945 (to GDP), MH-078113 (to MSK), MH-077862 (to JAS), MH-072767 (to SKH), and MH-103366 (to BAC), MH-103368 (to ESG).

## Author contributions

W.Y. conceptualized and designed the study. Data collection was done by C.A.T., E.S.G., B.A.C., G.D.P., S.S.K., S.K. H., J.E.M., M.S.K., and data analysis were performed by W.Y., B.K., and J.B.G. W.Y., B.K., and J.B.G. wrote the manuscript with input and critical revisions from C.A.T., E.C.DR., P.S., C.X., N. AR., E.S.G., B.A.C., G.D.P., S.S.K., E.I., S.K.H., J.E.M., M.S.K. All authors reviewed and approved the final manuscript.

## Data availability statement

The data is available at The National Institute of Mental Health Data Archive: https://nda.nih.gov/edit_collection.html?id=2165

## Competing Interests Statement

Dr. Clementz reports being on the KyNexis SAB and the B-SNIP Diagnostics, LLC Board of Managers.

Dr. McDowell reports serving on the B-SNIP Diagnostics, LLC Board of Managers

Dr. Keedy reports serving on the B-SNIP Diagnostics, LLC Board of Managers

Dr. Ivleva reports serving on the B-SNIP Diagnostics, LLC Board of Managers. She also served on scientific advisory boards for Janssen Scientific Affairs, LLC, Alkermes, Inc and Karuna Therapeutics.

Dr. Gershon reports serving on the B-SNIP Diagnostics, LLC Board of Managers

Dr. Keshavan reports serving on the B-SNIP Diagnostics, LLC Board of Managers

Dr. Pearlson reports serving on the B-SNIP Diagnostics, LLC Board of Managers

Dr. Tamminga reports serving on the Merck DSmB, being on the Karuna SAB and owning Karuna stock, and being on the KyNexis SAB and owning KyNexis stock, and B-SNIP Diagnostics, LLC Board of Managers.

Other authors have no competing interests to declare.

