## Supplementary Materials for "Exposotypes in Psychotic Disorders"

**Table S1:** Demographics of BSNIP2 (n = 1973).

|  | HC  (n = 623) | Proband  (n = 1350) | Total  (n = 1973) |  | | |
| --- | --- | --- | --- | --- | --- | --- |
|  | M (SD) | | | DF | *X*^2^/F | *p* |
| Age |  | | | 1 | 27.05 | < 0.001 |
|  | 34.37 (12.03) | 38.69 (18.99) | 37.32 (17.21) |  | | |
| Sex |  | | | 1 | 22.75 | < 0.001 |
| Male | 239 (38.4%) | 674 (50.1%) | 913 (46.4%) |  | | |
| Female | 383 (61.6%) | 672 (49.9%) | 1055 (53.6%) |  |  |  |
| Site |  | | | 4 | 12.67 | 0.013 |
| 2 | 56 (12.9%) | 115 (14.6%) | 171 (14.0%) |  | | |
| 3 | 100 (23.1%) | 200 (25.4%) | 300 (24.6%) |  |  |  |
| 4 | 58 (13.4%) | 112 (14.2%) | 170 (13.9%) |  |  |  |
| 6 | 108 (24.9%) | 130 (16.5%) | 238 (19.5%) |  |  |  |
| 7 | 111 (25.6%) | 229 (29.1%) | 340 (27.9%) |  |  |  |
| Diagnosis |  | | | — | | |
| SZ | – | 560 (41.5%) |  |  | | |
| SAD | – | 500 (37%) |  |  | | |
| BPD | – | 290 (21.5%) |  |  | | |

*BSNIP: Bipolar-Schizophrenia Network for Intermediate Phenotypes, HC: Healthy Control, Proband: Participant with psychosis-spectrum diagnosis, M: Mean, SD: Standard Deviation, DF: Degrees of Freedom, X*^2^*: Chi-square test statistic, F: Anova Test statistic, SZ: Schizophrenia, SAD: Schizoaffective Disorder, BPD: Bipolar Disorder with Psychotic Features.*

**Table S2:** Clustering performance outcomes across the Exposotype sample.

| Accuracy | Precision | Recall | F1-score |
| --- | --- | --- | --- |
| 92% | 92% | 92% | 92% |

**Table S3:** Clinical Outcomes and Comparisons for HC Sample

|  | Outcome | ET1 (◼) | ET2 (◻) | ET3 (⊗) | ET4 (Δ) |  | | | |
| --- | --- | --- | --- | --- | --- | --- | --- | --- | --- |
|  |  | M (SD) | | | | DF | F | *η^2^G* | *q* |
| Clinical | BIS-11 | 60.06 (11.63) | 58.36 (8.53) | 53.88 (9.04) | 52.04 (8.87) | 3 | 8.38 | 0.093 | ◼◻⊗Δ = ***  ◼⊗ = *  ◼Δ = **  ◻⊗ = *  ◻Δ = ** |
| Functioning | GAF | 82.47 (9.28) | 84.97 (4.21) | 85.33 (5.93) | 84.3 (7.16) | 3 | 1.32 | 0.017 | ◼◻⊗Δ = 0.27 |
|  | Birchwood: Social Functioning | 154.38 (20.14) | 149.21 (20.08) | 156.62 (18.03) | 152.02 (17.79) | 3 | 1.55 | 0.019 | ◼◻⊗Δ = 0.27 |
| Cognition | BACS | -0.401 (1.32) | -0.366 (1.19) | -0.063 (1.13) | -0.606 (1.12) | 3 | 3.223 | 0.039 | ◼◻⊗Δ = *  ⊗Δ = * |

*ET: Exposotype, ◼: ET1, ◻: ET2, ⊗: ET3, Δ:ET4, M: Mean, SD: Standard Deviation, DF: Degrees of Freedom, F: F-Statistic, FDR: False Discovery Rate Adjusted P-Values. BIS-11: Barratt Impulsiveness Scale, GAF: Global Assessment of Functioning, BACS: Brief Assessment of Cognition in Schizophrenia, *: p < 0.05, **: p < 0.01, ***: p < 0.001.*

**Table S4:** Exposotypes and differences in Estimated Total Intracranial Volume, Total Gray Matter Volume, and Hippocampus in the Psychosis Disorder Sample (n = 519).

| Outcome | ET1 (◼) | ET2 (◻) | ET3 (⊗) | ET4 (Δ) |  | | | |
| --- | --- | --- | --- | --- | --- | --- | --- | --- |
|  | M (SD) | | | | DF | F | *η^2^G* | *q* |
| Intracranial Volume | 1486218.124 (206393.523) | 1362566.488 (209490.605) | 1492342.918 (194438.433) | 1415342.104 (205121.695) | 3 | 8.95 | 0.067 | ◼◻⊗Δ = ***  ◼◻ = ***  ◻⊗ = ***  ⊗Δ = * |
| Total Gray Matter ↥ | 4188.258 (359.212) | 4295.909 (468.911) | 4194.148 (314.892) | 4250.258 (465.897) | 3 | 1.49 | 0.012 | ◼◻⊗Δ = 0.217 |
| Left Hippocampus ↥ | 23.917 (2.719) | 24.576 (3.523) | 23.813 (2.51) | 24.33 (3.193) | 3 | 1.32 | 0.01 | ◼◻⊗Δ = 0.268 |
| Right Hippocampus ↥ | 24.408 (2.82) | 25.038 (3.539) | 24.519 (2.417) | 24.96 (3.107) | 3 | 1.02 | 0.008 | ◼◻⊗Δ = 0.384 |

*ET: Exposotype, ◼: ET1, ◻: ET2, ⊗: ET3, Δ:ET4, M: Mean, SD: Standard Deviation, DF: Degrees of Freedom, F: F-Statistic, η^2^G: Generalized Eta Squared, q: False-discovery rate converted p-value. ↥: Adjusted for Intracranial Volume (ICV) by dividing the Region of Interest (RIO) by Intracranial Volume (ICV) (ROI/ICV), then multiplying by a factor of 10,000. For all pairwise testing: ***: q < 0.001, **: q < 0.01, * q < 0.05.*

**Figure S1:** Pairwise comparisons outcomes across Exposotypes for MRI Regions of Interest within the Psychosis Disorder Sample.

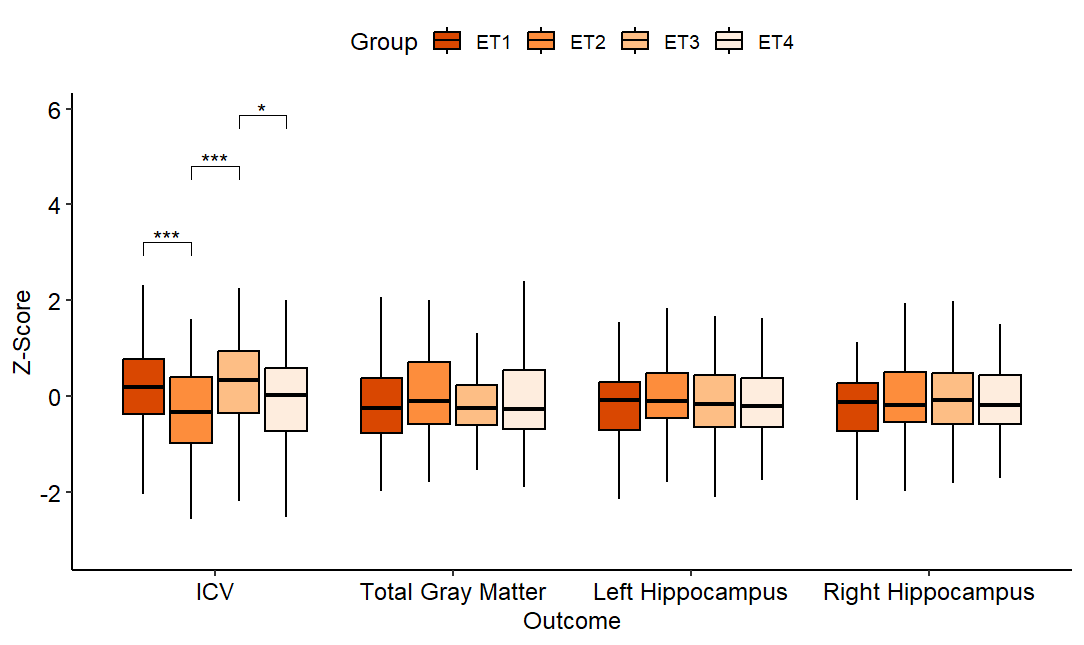

*ICV: Intracranial Volume, ET: Exposotype. ***: q < 0.001, **: q < 0.01, * q < 0.05.*

**Table S5:** Exposotypes and differences for MRI Regions of Interest within the Health Control Sample (n = 253).

| Outcome | ET1 (◼) | ET2 (◻) | ET3 (⊗) | ET4 (Δ) |  | | | |
| --- | --- | --- | --- | --- | --- | --- | --- | --- |
|  | M (SD) | | | | DF | F | *η^2^G* | *q* |
| Intracranial Volume | 1461019.535 (140907.281) | 1433678.097 (141417.75) | 1520056.259 (166167.447) | 1428009.107 (173832.065) | 3 | 4.32 | 0.062 | ◼◻⊗Δ = **  ◻⊗ = *  ⊗Δ = ** |
| Total Gray Matter Volume↥ | 4364.081 (251.071) | 4373.963 (341.278) | 4302.645 (334.241) | 4363.224 (374.837) | 3 | 0.51 | 0.008 | ◼◻⊗Δ = 0.673 |
| Left Hippocampus ↥ | 24.596 (2.333) | 24.354 (2.429) | 24.015 (2.3) | 24.973 (2.534) | 3 | 1.98 | 0.029 | ◼◻⊗Δ = 0.118 |
| Right Hippocampus ↥ | 25.068 (2.742) | 25.006 (2.517) | 24.14 (2.244) | 25.454 (2.657) | 3 | 3.44 | 0.05 | ◼◻⊗Δ = *  ⊗Δ = ** |

*ET: Exposotype, ◼: ET1, ◻: ET2, ⊗: ET3, Δ:ET4, M: Mean, SD: Standard Deviation, DF: Degrees of Freedom, F: F-Statistic, η^2^G: Generalized Eta Squared, q: False-discovery rate converted p-value. ↥: Adjusted for Intracranial Volume (ICV) by dividing the Region of Interest (RIO) by Intracranial Volume (ICV) (ROI/ICV), then multiplying by a factor of 10,000. For all pairwise testing: ***: q < 0.001, **: q < 0.01, * q < 0.05.*

**Figure S2:** Pairwise comparisons outcomes across Exposotypes for MRI Regions of Interest within the Healthy Control Sample.

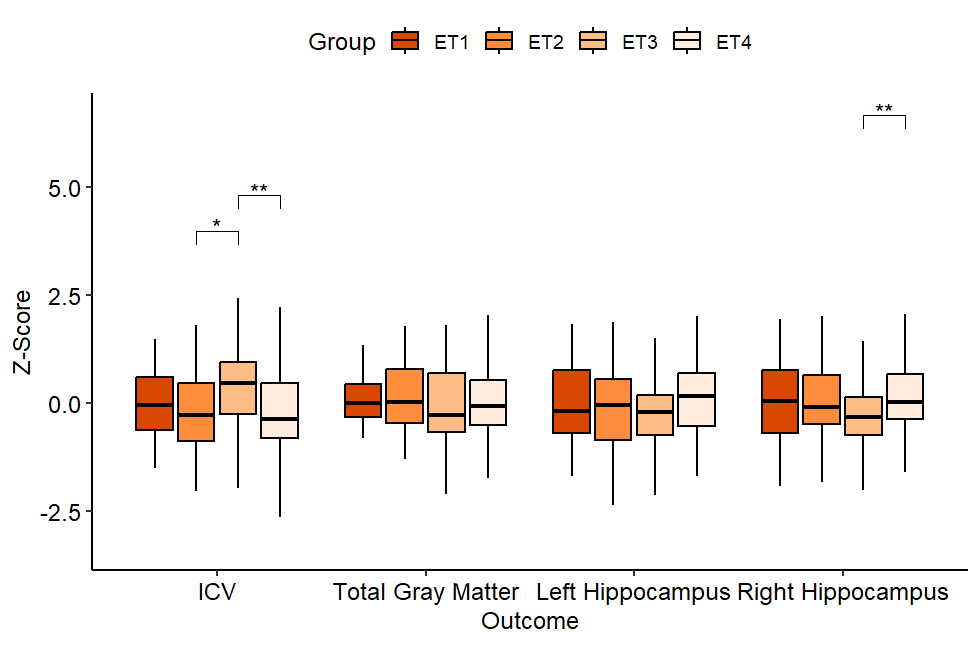

*ICV: Intracranial Volume, ET: Exposotype. ***: q < 0.001, **: q < 0.01, * q < 0.05.*

**Figure S3:** Exposotype prevalence among DSM-V and Biotypes diagnostic categories

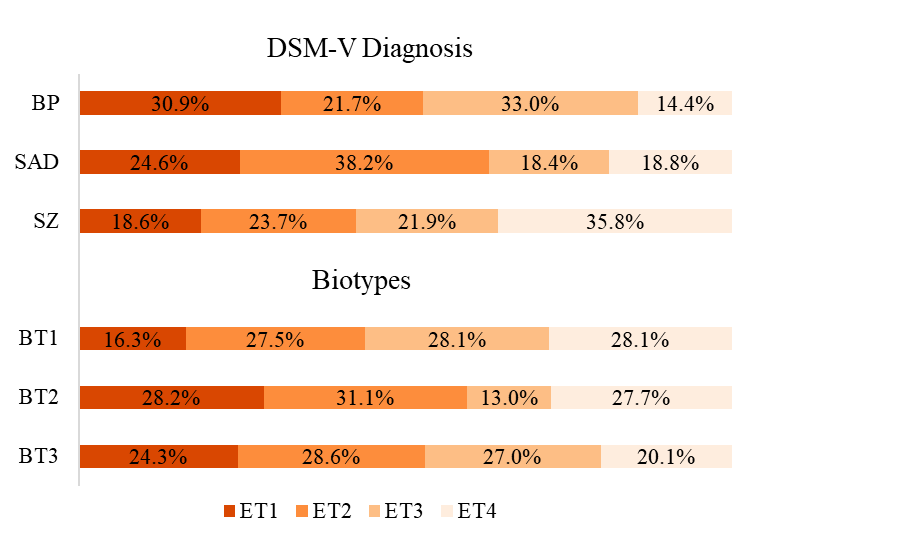

*BP: Bipolar disorder with psychotic features, SAD: Schizoaffective Disorder, SZ: Schizophrenia, BT: Biotype, ET: Exposotype, DSM-V: Diagnostic and Statistical Manual Version 5.*

**Table S6:** Control distribution in ETs

| Outcome | ET1 (◼) | ET2 (◻) | ET3 (⊗) | ET4 (Δ) |
| --- | --- | --- | --- | --- |
| C | 31 (12.25%) | 35 (13.83%) | 77 (30.43%) | 110 (43.48%) |

*ET: Exposotype, ◼: ET1, ◻: ET2, ⊗: ET3, Δ:ET4, C: Control.*
